# Core warming of coronavirus disease 2019 (COVID-19) patients undergoing mechanical ventilation - a pilot study

**DOI:** 10.1101/2023.04.12.23287815

**Authors:** Nathaniel P Bonfanti, Nicholas M Mohr, David C Willms, Roger J Bedimo, Emily Gundert, Kristina L Goff, Erik B Kulstad, Anne M Drewry

## Abstract

Fever is a recognized protective factor in patients with sepsis, and growing data suggest beneficial effects on outcomes in sepsis with elevated temperature, with a recent pilot randomized controlled trial showing lower mortality by warming afebrile sepsis patients in the intensive care unit. The objective of this prospective single-site randomized controlled trial was to determine if core warming improves respiratory physiology of mechanically ventilated patients with COVID-19, allowing earlier weaning from ventilation, and greater overall survival. A total of 19 patients with mean age of 60.5 (±12.5) years, 37% female, mean weight 95.1 (±18.6) kg, and mean BMI 34.5 (±5.9) kg/m2 with COVID-19 requiring mechanical ventilation were enrolled from September 2020 through February 2022. Patients were randomized 1:1 to standard-of-care or to receive core warming for 72 hours via an esophageal heat exchanger commonly utilized in critical care and surgical patients. The maximum target temperature was 39.8 °C. A total of 10 patients received usual care and 9 patients received esophageal core warming. After 72 hours of warming, PaO2/FiO2 ratios were 197 (±32) and 134 (±13.4), Cycle Thresholds were 30.8 (±6.4) and 31.4 (±3.2), ICU mortality was 40% and 44%, 30-day mortality was 30% and 22%, and mean 30-day ventilator-free days were 11.9 (±12.6) and 6.8 (±10.2) for standard-of-care and warmed patients, respectively (p=NS). This pilot study suggests that core warming of patients with COVID-19 undergoing mechanical ventilation is feasible and appears safe. Optimizing time to achieve febrile-range temperature may require a multimodal temperature management strategy to further evaluate effects on outcome.

## Background

Fever is a recognized protective factor in patients with sepsis (Rumbus et al., 2017). Prospective data have shown that afebrile patients have higher 28-day mortality (37.5% vs 18.2%), increased acquisition of secondary infections (35.4% vs. 15.9%) and suppressed HLA-DR expression suggestive of monocyte dysfunction over time (Drewry et al., 2018). Elevated temperatures have been shown to augment immune function, increase production of protective heat shock proteins, directly inhibit microorganism growth, reduce viral replication, and enhance antibiotic effectiveness (Drewry and Hotchkiss, 2013, Launey et al., 2011). The first pilot randomized controlled trial (RCT) of warming as a therapy for sepsis found lower mortality by warming afebrile patients in the intensive care unit (ICU) (Drewry et al., 2022). Conversely, randomized controlled trials have failed to find benefits to reducing fever of infectious etiology (Dallimore et al., 2018, Drewry et al., 2017, Gozzoli et al., 2001, Peters et al., 2019, Schulman et al., 2005, Young et al., 2015, Young et al., 2019, Zhang, 2015), and therapeutic hypothermia in sepsis has been found to be either of no benefit, or harmful (Itenov et al., 2018, Saoraya et al., 2020).

Innate and adaptive immunological processes appear to be accelerated by fever (Evans et al., 2015, Lee et al., 2012, Peters et al., 2019). More rapid recovery from chickenpox (Doran et al., 1989), malaria (Brandts et al., 1997), and rhinovirus (Stanley et al., 1975) infections have been shown by avoiding antipyretic medication. Because many viruses replicate more robustly at cooler temperatures, such as those found in the nasal cavity (33–35°C) than at warmer core body temperature (37°C) (Chan et al., 2011, Foxman et al., 2015, Laporte et al., 2019, Ping, 2003, Zou et al., 2020), elevated temperature may offer another benefit in treating viral illnesses. The virus causing coronavirus disease 2019 (COVID-19), SARS-CoV-2 may behave similarly to other viruses susceptible to temperature changes (Wang et al., 2020). The fact that fever has often abated by the time a COVID-19 patient requires mechanical ventilation may offer a specific window of opportunity for treatment with therapeutic hyperthermia (Arabi et al., 2022, Chan et al., 2011, Foxman et al., 2015, Roger, 2021, Zhou et al., 2020).

The first RCT of warming therapy in sepsis found that by using forced-air warming, not all patients were able to achieve target temperature (Drewry et al., 2022). Additional means of providing heat to patients may therefore be desirable. The provision of heat at the patient’s core (where viral replication may be greatest) rather than via peripheral heat transfer across the skin may offer further mechanistic benefits. A dedicated device (ensoETM, Attune Medical, Chicago, IL) offers a means to provide heat through the esophagus using a closed-loop flow of water that can be adjusted to cause temperature change (Furrer et al., 2022). Toward this goal, the aim of this pilot study was to investigate the feasibility of providing febrile-range temperatures to COVID-19 patients requiring mechanical ventilation via core warming, in order to determine if core warming improves respiratory physiology of mechanically ventilated patients with COVID-19, allowing earlier weaning from ventilation, and greater overall survival.

## Materials and Methods

This was a single-center randomized controlled pilot trial (NCT04494867), and we have previously published the study protocol (Bonfanti et al., 2020). Participants were included if they were age 18 or older, diagnosed with COVID-19, intubated for respiratory failure, and had a maximum baseline temperature of < 38.3 °C. We excluded patients with a contraindication to core warming using an esophageal core warming device, pregnancy, body weight less than 40 kg, a Do-Not-Resuscitate (DNR) order, history of acute stroke, post-cardiac arrest, or multiple sclerosis. Written, informed consent was obtained from each patient’s legally authorized representative prior to enrollment. All procedures followed were in accordance with the ethical standards of the Sharp Memorial Hospital IRB (IRB # 2007901; approved August 17, 2020) and with the Helsinki Declaration of 1975.

The primary outcome was the severity of acute respiratory distress syndrome as measured by PaO2/FiO2 ratio. Secondary outcomes included the change in viral load measured in lower respiratory tract samples, the duration of mechanical ventilation, and mortality. Patient core warming was provided by a commercially available esophageal heat exchange system (ensoETM, Attune Medical, Chicago, IL). The device was set to 42°C after initiation and maintained at 42°C for the duration of 72 hours of treatment unless patient temperature exceeded 39.8°C, at which point the device setpoint was reduced to 40°C. Control patients were provided standard temperature management.

## Results

A total of 19 patients were randomized (10 patients to control, 9 to core warming). The patients had a mean age of 60.5 (±12.5) years, 37% were female, with a mean weight of 95.1 (±18.6) kg, and a mean BMI 34.5 (±5.9) kg/m2. Patient baseline demographics by group are as shown in Table 1.

**Table 1.**
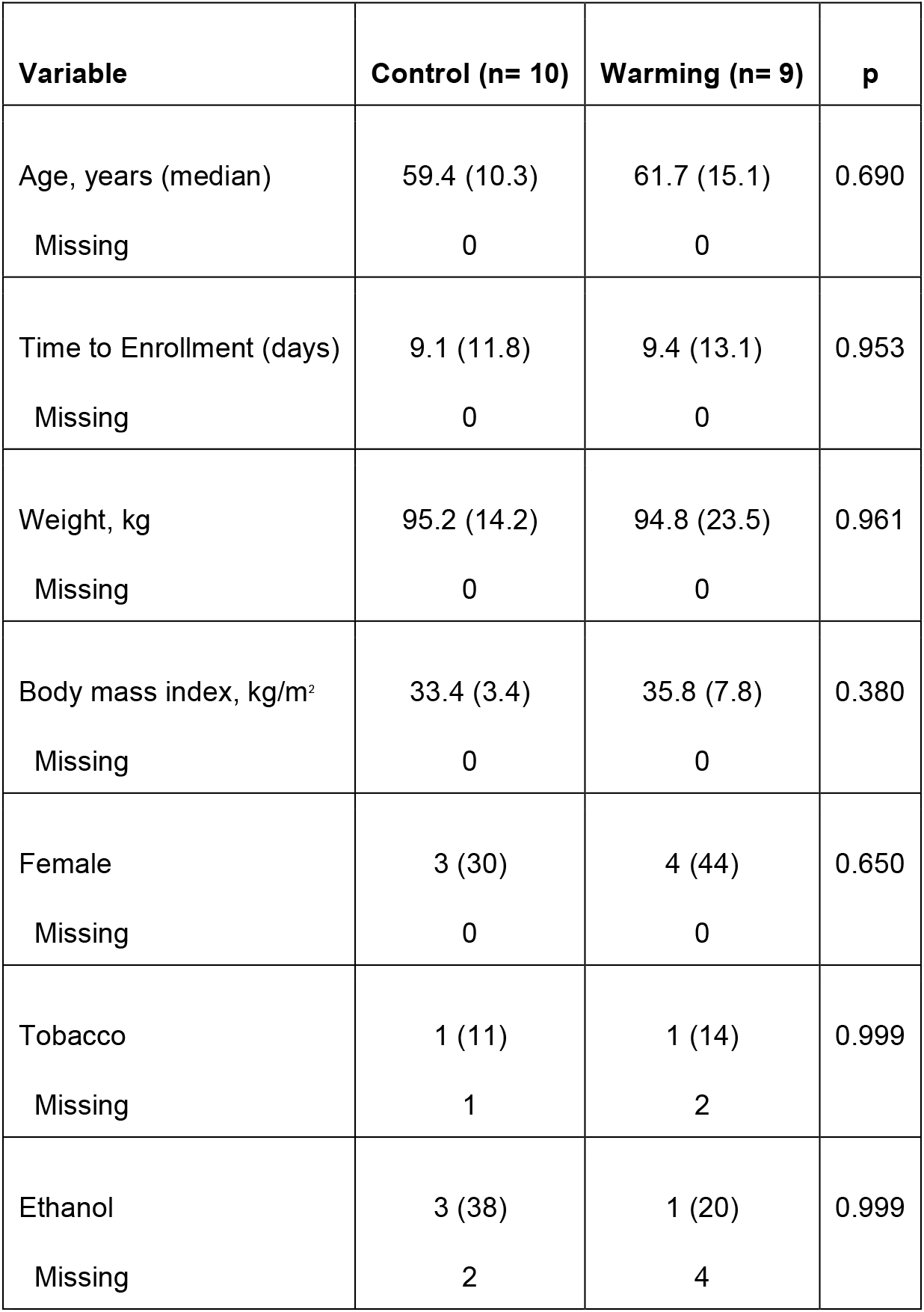
Baseline demographics.

| Variable | Control (n= 10) | Warming (n= 9) | p |
| --- | --- | --- | --- |
| Age, years (median) | 59.4 (10.3) | 61.7 (15.1) | 0.690 |
| Missing | 0 | 0 |  |
| Time to Enrollment (days) | 9.1 (11.8) | 9.4 (13.1) | 0.953 |
| Missing | 0 | 0 |  |
| Weight, kg | 95.2 (14.2) | 94.8 (23.5) | 0.961 |
| Missing | 0 | 0 |  |
| Body mass index, kg/m<sup>2</sup> | 33.4 (3.4) | 35.8 (7.8) | 0.380 |
| Missing | 0 | 0 |  |
| Female | 3 (30) | 4 (44) | 0.650 |
| Missing | 0 | 0 |  |
| Tobacco | 1 (11) | 1 (14) | 0.999 |
| Missing | 1 | 2 |  |
| Ethanol | 3 (38) | 1 (20) | 0.999 |
| Missing | 2 | 4 |  |

Temperature curves for each patient group are shown in Figure 1. Good separation between groups was obtained.

**Figure 1.**
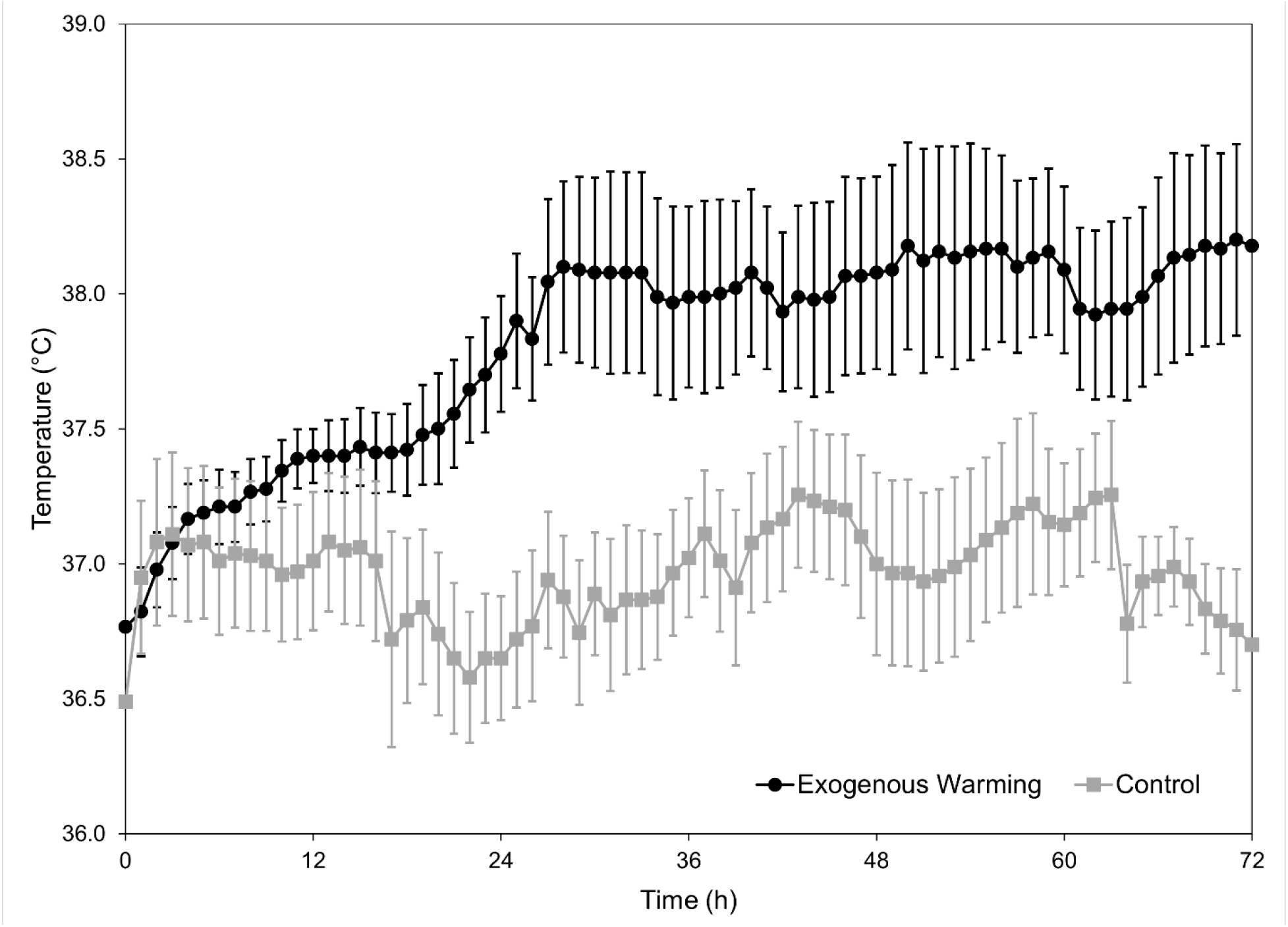
Patient temperature curves over 72 hours of treatment.

Measured outcomes are shown in Table 2. The outcomes were similar in this small pilot study. There was a single adverse event attributed to the use of the device, reported as an episode of nausea, vomiting, and abdominal pain.

**Table 2.** Outcomes.

| Variable | Control (n=10) | Warming (n=9) | P |
| --- | --- | --- | --- |
| Cycle threshold, 72 h | 30.8 (6.4) | 31.4 (3.2) | 0.852 |
| Missing | 3 | 3 |  |
| Delta-cycle threshold | 4.4 (1.4) | 0.2 (2.0) | 0.115 |
| Missing | 4 | 4 |  |
| ICU Mortality | 4 (40) | 4 (44) | 0.999 |
|  | 0 | 0 |  |
| 30-day Mortality | 3 (30) | 2 (22) | 0.999 |
|  | 0 | 0 |  |
| PaO<sub>2</sub>/FiO<sub>2</sub> at t=0 | 165 (19.4) | 119 (11.6) |  |
| PaO<sub>2</sub>/FiO<sub>2</sub> at t=24 hours | 155 (21.2) | 130 (17.0) |  |
| PaO<sub>2</sub>/FiO<sub>2</sub> at t=48 hours | 197 (30.6) | 158 (19.4) |  |
| PaO<sub>2</sub>/FiO<sub>2</sub> at t=72 hours | 197 (32.0) | 134 (13.4) |  |

## Discussion

Growing data support beneficial effects from warming patients with severe infections, and there are increasing research efforts being undertaken to better understand the mechanistic underpinnings for the clinical effects reported. Challenges in warming patients to febrile-range hyperthermia will need to be adequately addressed in order to fully explore hyperthermia as a treatment. This pilot study suggests that core warming provided by a commercially available esophageal heat transfer system appears safe and can provide febrile-range temperatures to patients with COVID-19 undergoing mechanical ventilation.

Core body temperature is far more tightly regulated than other physiological parameters (including blood pressure and heart rate), and because of the existence of robust thermal defense mechanisms, warming patients to above normal temperatures can be difficult (Sessler, 2009). Previous data have found that core warming with a dedicated esophageal heat transfer device is able to provide warming in patients with particular difficulty in maintaining normothermia, such as burn patients undergoing surgery (Furrer et al., 2022). At present, it remains unclear if a shorter time to a febrile target temperature is beneficial or will be required to demonstrate benefits seen in early clinical studies.

Further studies are in development which aim to investigate further mechanistic underpinnings behind the effects of elevated temperature in severe infections. Given the known challenges in warming patients to above-normal body temperature, a multimodal temperature management strategy may be required, and is being anticipated, in subsequent investigations.

Limitations of this study include the small sample size, the inability to blind healthcare providers treating enrolled patients, and the lack of measurement of specific immune factors such as IL-1, IL-6, tumor necrosis factor (TNF), and interferon (IFN).

## Conclusions

Core warming of patients with COVID-19 undergoing mechanical ventilation is feasible and appears safe. To optimize time to achieve febrile-range temperature in subsequent studies, a multimodal temperature management strategy may be necessary.

## Data Availability

All data produced in the present study are available upon reasonable request to the authors.

## Acknowledgements

The investigators thank DeAnn S. Cary, Ph.D, MacKenzie Habib, BS, Kyra Rashid, BS, MCR, Kathryn Hong, Cary Murphy, RN, Christine Kappel, RN, the Sharp Memorial Hospital, and the Sharp Center for Research.

## Contributions

Dr. Bonfanti: Writing – original draft, review, and editing.

Dr. Mohr: Methodology, data curation, and formal analysis

Dr. Willms: Project administration, resources, investigation

Dr. Bedimo, MD: Conceptualization and methodology

Dr. Gundert, MD: Conceptualization

Dr. Goff, MD: Conceptualization

Dr. Kulstad, MD, MS: Conceptualization, methodology, project administration, writing - review and editing.

Dr. Drewry, MD, MS: Conceptualization and methodology

